# Clinical Effectiveness of cardiac rehabilitation utilisation and the barriers to its completion among patients of low socioeconomic status living in rural areas: a mixed methods study

**DOI:** 10.1101/2023.11.09.23298047

**Authors:** Alline Beleigoli, Hila Ariela Dafny, Maria Alejandra Pinero de Plaza, Claire Hutchinson, Tania Marin, Joyce S Ramos, Orathai Suebkinorn, Lemlem G. Gebremichael, Norma B. Bulamu, Wendy Keech, Marie Ludlow, Jeroen Hendriks, Vincent Versace, Robyn A. Clark the Country Heart Attack Prevention (CHAP) Project team

## Abstract

**Background:** Low socioeconomic status (LSES) and rurality are associated with poor cardiovascular outcomes and reduced cardiac rehabilitation (CR) participation.

**Aim:** To investigate CR utilization and effectiveness, factors, needs and barriers associated with non-completion among patients of LSES in rural Australia.

**Methods:** Through a concurrent triangulation mixed methods design we converged the results of a retrospective cohort and a qualitative study. A Cox survival model applied to a population balanced by inverse probability weighting assessed the association between CR utilization and 12-month mortality/cardiovascular readmissions. Associations with non-completion were tested by logistic regression. Barriers and needs to CR completion were evaluated by thematic analysis of semi-structured interviews and focus groups with 28 participants.

**Results:** Among 16,159 eligible separations, 44.3% were referred and 11.2% completed CR. Completing CR (HR 0.65; 95%CI 0.57-0.74; p<0.001) led to a lower risk of cardiovascular readmission/death. Living alone (OR 1.38; 95%CI 1.00-1.89; p=0.048), having diabetes (OR 1.48; 95%CI 1.02-2.13; p=0.037), or having depression (OR 1.54; 95%CI 1.14-2.08; p=0.005), were associated with a higher risk of non-completion whereas enrolment in a telehealth program was associated with a lower risk of non-completion (OR 0.26; 95%CI 0.18-0.38; p<0.001). Themes related to logistic issues, social support, transition of care challenges, lack of care integration, and of person-centeredness emerged as barriers to CR completion.

**Conclusions:** CR completion was low but effective in reducing mortality/cardiovascular readmissions. Understanding and addressing barriers and needs through mixed methods can help tailor CR programs to vulnerable populations and improve completion and outcomes.

## Introduction

Low socioeconomic status is associated with poor cardiovascular outcomes.^1^ Compared to high socioeconomic areas, the cardiovascular death rate in low socioeconomic areas is 1.3 times higher for males and 1.5 times higher for females.^2^ Economic stability, neighborhood-built environment, education and health care access, social and community relationships can be sources of chronic psychosocial stress. These stressors can activate biological responses that lead to autonomic dysfunction and chronic inflammation.^3^ All these processes contribute to an increased cardiovascular risk and explain why psychosocial risk factors (e.g., low socioeconomic status, low social support, depression, anxiety) tend to cluster together and co-occur with traditional risk factors (e.g., smoking, obesity, diabetes), ultimately resulting in increased cardiovascular events and mortality.^3^

These social determinants of health can be more prominent in rural than in metropolitan areas. Socio-economic status is typically lower in rural and remote areas relative to metropolitan areas.^4^ This, in combination with worse health services access in rural areas, may contribute to poorer cardiovascular outcomes reported for these areas. In Australia, people living in remote areas have a 1.4 times higher age-adjusted hospitalisation rate than those living in major cities. For coronary heart disease, the leading cause of death, age-adjusted death rates are 1.3 times higher in remote areas than Australia overall. ^2^

Cardiac rehabilitation can improve functional capacity and traditional cardiovascular risk factors, accounting for much of the association between low socioeconomic status and cardiovascular mortality.^5^ Cardiac rehabilitation is guideline-recommended, ^6, 7^ and effective in reducing mortality and hospital readmission at 12 months post-discharge. ^8, 9^ However, globally only 30-50% of eligible patients are referred, around 30% of the referred attend at least one session, and 10-30% complete the program.^10^

Patients living in low-income areas are less likely to attend or complete cardiac rehabilitation after hospitalisation for a heart attack, and to be informed about cardiac rehabilitation while in hospital.^11, 12^ We aimed to evaluate cardiac rehabilitation utilisation and effectiveness on clinical outcomes (readmissions and overall deaths) of patients with low socioeconomic status living in rural South Australia. Additionally, we sought to understand barriers to cardiac rehabilitation completion and the needs of patients with low socioeconomic status residing in rural areas.

## Methods

### Study design

This mixed-methods study has a concurrent triangulation and convergent parallel design.^13^ The quantitative analysis used a retrospective cohort design, including patients admitted for a cardiovascular event from 01/01/2016 to 30/06/2021. The qualitative study involved four semi-structured interviews and five focus groups with participants from low socioeconomic status areas in rural South Australia. Factors associated with non-completion and reasons for non-completion retrieved from the quantitative study will be compared and related to the themes and subthemes derived from the thematic analysis of the qualitative study ^13^ (Figure 1).

**Figure 1:**
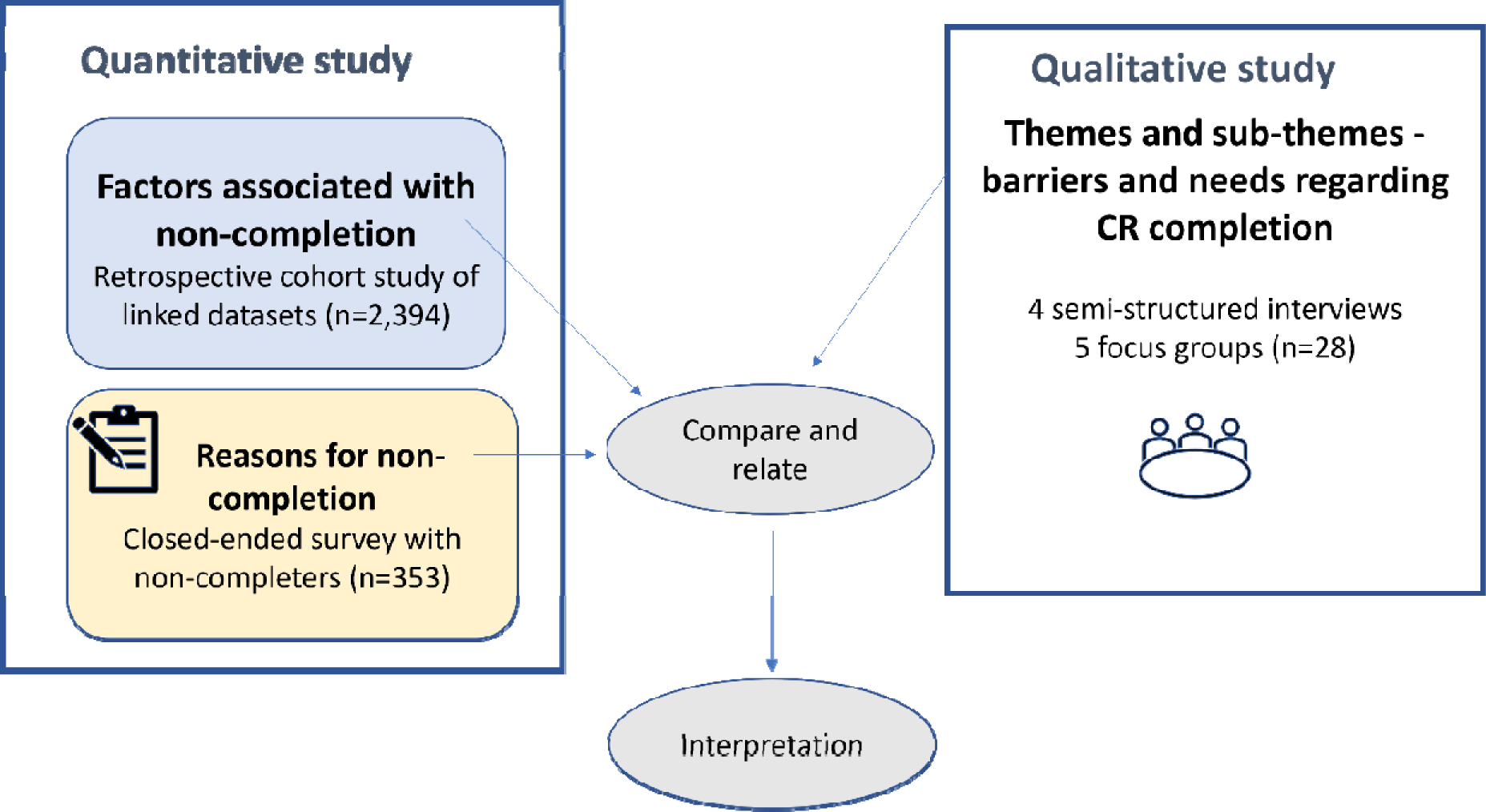
Mixed-methods study diagram.

### Definitions

Low socioeconomic status was defined as an Index of Relative Socioeconomic Advantage and Disadvantage score within the 2 lowest deciles. The Index of Relative Socioeconomic Advantage and Disadvantage measures high and low income, degree of house mortgage, size of home, educational level, qualifications or none, employed as a professional, a manager, low skilled worker, machinery operator or laborer, high rent, number of cars or none, overcrowding, divorced, low rent, disability, unemployed, single parent family, no internet access and jobless parents from the Australian Census.^14^

Rural areas were defined as those outside metropolitan areas according to the Modified Monash Model which also consider distance to services to classify rural (also named regional in Australia) according to its remoteness (inner regional; outer regional; remote and very remote areas).^4^

### Setting

All South Australian public hospitals and the 15 public cardiac rehabilitation programs in rural areas, including the one delivered exclusively via telephone, were involved. All cardiac rehabilitation programs receive referrals from metropolitan and regional hospitals via a central referral system. All programs comprised the following core components: referral and access to services; clinical assessment and short-term monitoring; recovery and long-term maintenance; lifestyle/behavioral modification and medication adherence; and evaluation and quality improvement.^15^

### Quantitative study

#### Participants

Eligible patients were adults (≥18 years); living in rural areas of South Australia with a low socioeconomic status; discharged alive from a hospital separation with a primary or secondary cardiac-related diagnostic code, with or without an interventional procedure, including acute coronary syndrome, heart failure, atrial fibrillation, acute myocardial infarction, percutaneous coronary intervention, coronary artery bypass graft, aortic or mitral valve replacements, automatic implantable cardioverter-defibrillator or permanent pacemaker.

A hospital separation was defined as from the beginning of admission to discharge. Separations across multiple hospitals, transfers, and readmissions within 24 hours were merged into one event.

#### Variables/outcomes

The primary outcome was cardiac rehabilitation utilisation measured across 3 categories: did not receive cardiac rehabilitation (not referred or declined), commenced did not complete cardiac rehabilitation, and completed cardiac rehabilitation. Referrals were defined by a health professional assessment and recorded in the South Australian cardiac rehabilitation program database. Commencement (≥1 session) and completion as defined by a clinician were also characterised by a health professional involved in the patient care.

Secondary outcomes included a composite endpoint of all-cause mortality or admission for new/re-myocardial infarction, heart failure, atrial fibrillation, or stroke within 12 months of the index admission.

#### Data sources/measurements and linkage process

The public hospital’s database (South Australian Admitted Patient Care), the Emergency Department Data Collection, the South Australian Births, Death and Marriages, and the South Australian cardiac rehabilitation datasets were linked through deterministic linkages based on 11 identifying variables (client identification number, last name, first name, other names, date of birth, sex, date of death, weight at birth, street address, suburb, and postcode). Records not linked deterministically were analysed by probabilistic linkage and subsequently underwent clerical review if still unmatched.

The South Australian cardiac rehabilitation database captured dates of referrals, commencements, and completions; pre, post, 6 and 12-month cardiovascular risk factors, depression screening through Patient Health Questionnaire (PHQ)-2 and PHQ-9;^16^ self-reported smoking and medication adherence; and exercise capacity. Additionally, it captures a close-ended questions with a list of possible reasons for non-completion prompted by the clinicians to patients commencing but not completing cardiac rehabilitation.

#### Bias

We used an inverse probability weighting model adjusted for age, gender, primary diagnosis, Charlson Comorbidity Index,^17^ prior heart failure, coronary disease, atrial fibrillation, revascularisation, malignancy, and socioeconomic disadvantage measured by the Index of Relative Socioeconomic Advantage and Disadvantage^14^ to account for selection bias and unequal probabilities of cardiac rehabilitation completion. The balance of the inverse probability weighting model was tested by comparing the standardised difference (<10%) of the probabilities between the group that did not receive cardiac rehabilitation (was not referred or declined cardiac rehabilitation) vs. those who commenced but did not complete and those who completed cardiac rehabilitation (Supplementary Table 1). As a prior diagnosis of dementia was not balanced across the groups in the weighted model (i.e., standardised difference≥10%), we excluded patients with dementia (n=135).

#### Study size

The quantitative study used a convenience sample selecting all hospital records and linking all the separations eligible to cardiac rehabilitation from the South Australian Department of Health Admitted Patient Care database between January 2016 and June 2021 (5.5 years).

#### Quantitative variables and Statistical methods

We compared patient demographics, clinical, and social characteristics across the cardiac rehabilitation utilisation categories through Kruskal-Wallis (for continuous variables described by median and interquartile range) and Chi-square tests (for categorical variables defined by proportions). Cardiac rehabilitation utilisation was reported as ratios, measured as proportions (%).

To assess the association of the secondary outcome (composite of cardiovascular readmission due to myocardial infarction, heart failure, atrial fibrillation, or stroke and all-cause mortality) and cardiac rehabilitation utilisation (3 groups), we performed Cox proportional hazard models applied to the weighted population after testing for the proportional hazard assumption. The models were adjusted by age, gender, remoteness, primary cardiac diagnosis, referring hospital, and the Charlson Comorbidity Index.^17^ A p-value of <0.05 was statistically significant, and Stata 15.0 was used for the analysis.^18^

For the sub-cohort of separations receiving cardiac rehabilitation, we investigated the association of non-completion (independent variable) with sociodemographic and clinical factors by adjusting binary logistic models for age, gender, remoteness (inner regional, outer regional, remote, and very remote), living alone, English as a second language, index separation characteristics pre or during the COVID-19 pandemic, the reason for referral (acute myocardial infarction, heart failure, atrial fibrillation, coronary artery bypass graft, percutaneous coronary intervention, Valve procedures, Implantable valve procedures), Charlson Comorbidity Index, and history of comorbidities (diabetes, obesity, hypertension, depression) and the waiting time to commence cardiac rehabilitation from hospital separation.

### Qualitative study

#### Study design and data collection

Qualitative data from 28 participants were collected through four semi-structured interviews and five focus groups, using the Experience Based Co-Design – A toolkit for Australia^19^ as a framework to engage with the participants through accessible, tangible ways (e.g., visual communication, scenarios, and personas). This approach helps patients with low literacy levels convey ideas and design better service experiences.^19^ Perceived barriers and needs from hospital discharge/referral to (non) attendance/completion of cardiac rehabilitation were explored through interviews and focus groups. The research questions and target groups in this phase were specific, and the sample size generated sufficient information for an in-depth analysis of the narratives.^20^

#### Participants and recruitment

The cardiac rehabilitation nurse recruited participants; with inclusion criteria being adults (≥18 years), referred to a cardiac rehabilitation program in rural South Australia, living in an area of low socioeconomic status. Cognitive impairment (Mini-mental short scale<20) was an exclusion criterion.

#### Qualitative variables and data analysis

Interviews and focus groups interviews audio recordings were transcribed verbatim by a professional transcription company. The de-identified transcripts were analysed using Braun and Clark inductive thematic analysis.^21^ Familiarisation with the topic was established through individual organic and subjective coding by AB and HD. Nodes were established independently and then examined collaboratively by AB, HD, CH, and MAPP after identifying recurrent themes to confirm final themes. Verbatim quotes were extracted to illustrate themes and subthemes.

#### Bias

Reporting bias was mitigated by involving researchers from different disciplines and backgrounds (medicine, nursing, health economics, and public health) in the analysis. Additionally, these researchers were not involved in data collection. To reduce selection bias, we offered options for out-of-hours participation, and in-person or virtual/phone attendance. Participants were reimbursed for travel and participation time.

### Ethics

The study received approval from the South Australian Department for Health and Wellbeing Human Research Ethics Committee (2021/HRE00270) and the Southern Adelaide Clinical Human Research Ethics Committee (LNR/22/SAC/71). A waiver of consent was granted to the project by the Department for Health and Wellbeing Ethics Committee.

## Results

### Quantitative study participant characteristics

There were 16,159 separations eligible to this analysis after exclusion of patients with dementia (Figure 2). Participants’ mean age was 68.9 (SD 14.3) years, and 40.9% (n=6,610) were female, acute myocardial infarction (n= 1,773; 11.0%) and percutaneous coronary intervention (n=806; 4.9%) were the most frequent index admission single diagnosis and procedure, respectively. A referral was received by 7,159/16,159 (44.3%). Among the 7,159 who received a referral, 2,394 (33.4%) commenced cardiac rehabilitation and among those commencing, 1,806 (74.6%) completed the program. When assessed among the eligible, the completion rate was 11.2%. Characteristics of patients across the cardiac rehabilitation utilisation groups are displayed in Table 1.

**Figure 2:**
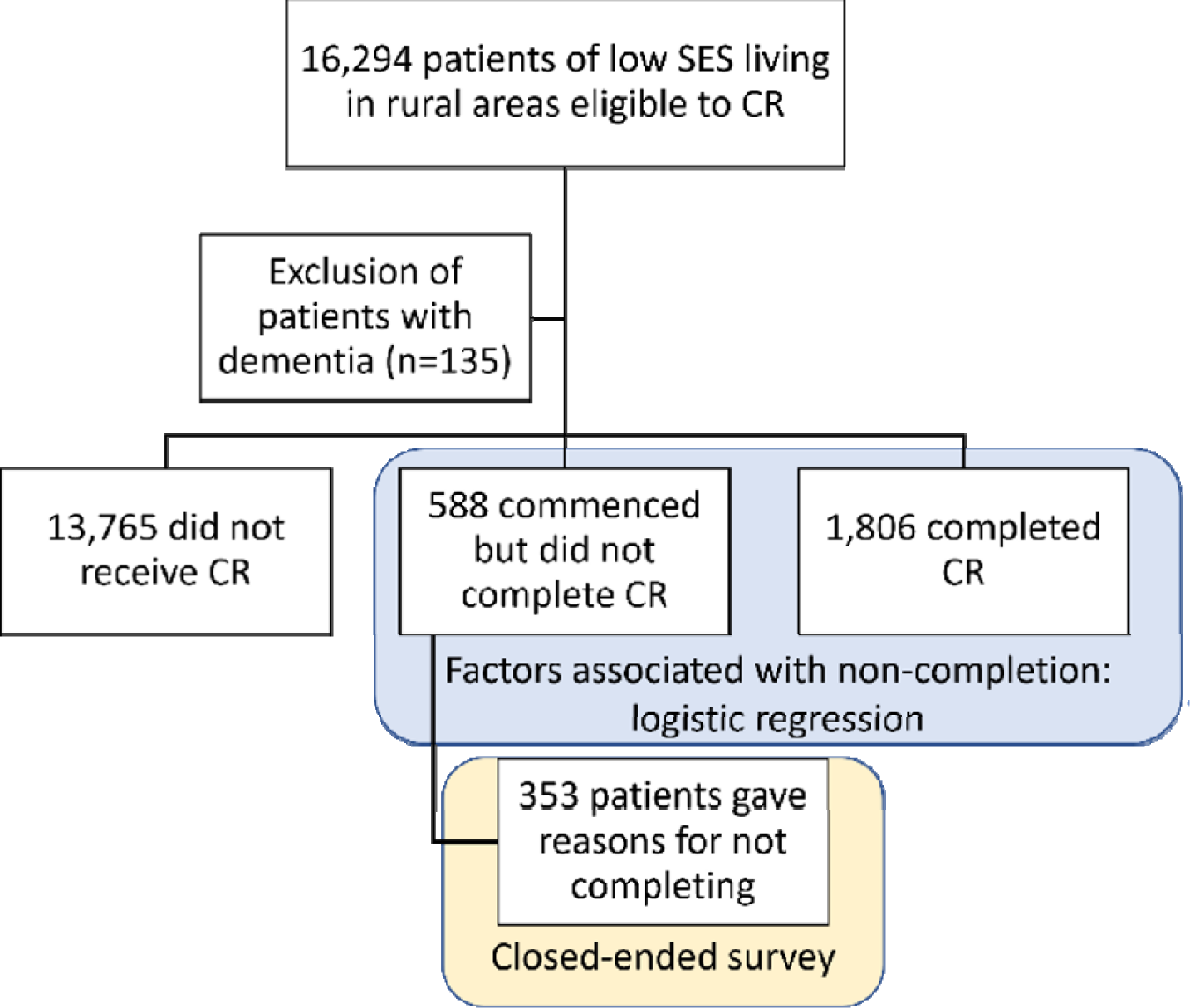
Quantitative study flow diagram.

**Table 1:** Characteristics of the patients across groups of cardiac rehabilitation utilisation for the non-weighted population.

| Characteristic | Overall<br>population<br>(16,159) | Did not receive<br>cardiac<br>rehabilitation<br>(n=13,765) | Commenced but did<br>not complete cardiac<br>rehabilitation<br>(n=588) | Completed cardiac<br>rehabilitation<br>(n=1,806) | p value |
| --- | --- | --- | --- | --- | --- |
| Age | 71 (60-79) | 71 (60-80) | 66 (57-73) | 68 (59-74) | 0.001 |
| Female Sex | 6610 (40.9) | 5879 (42.7) | 190 (32.3) | 541 (30.0) | <0.001 |
| Charlson Score | 1 (0-1) | 1 (0-1) | 1 (0-1) | 1 (0-1) | 0.286 |
| Separation during the<br>COVID pandemic | 1814 (11.2) | 1532 (11.1) | 57 (9.7) | 225 (12.5) | 0.118 |
| **Diagnosis at index**<br>**separation** |  |  |  |  |  |
| Acute MI | 1773 (11.0) | 1210 (8.8) | 124 (21.1) | 439 (24.3) | <0.001 |
| Congestive HF | 1501 (9.3) | 1373 (10.0) | 37 (6.3) | 91 (5.0) | <0.001 |
| Arrhythmia | 1520 (9.4) | 1368 (10.0) | 34 (5.8) | 118 (6.5) | <0.001 |
| **Procedures at index**<br>**separation** |  |  |  |  |  |
| CABG | 255 (1.6) | 194 (1.4) | 14 (2.4) | 47 (1.6) | <0.001 |
| PCI | 806 (5.0) | 516 (3.8) | 56 (9.5) | 234 (13.0) | <0.001 |
| Valve procedures | 508 (3.1) | 453 (3.3) | 12 (2.0) | 43 (2.4) | 0.034 |
| Implantable devices | 334 (2.1) | 302 (2.2) | 8 (1.4) | 24 (1.3) | 0.025 |
| **Additional**<br>**cardiovascular**<br>**diagnoses** |  |  |  |  |  |
| Prior MI | 1568 (9.7) | 1035(7.5) | 117 (19.9) | 416 (23.0) | <0.001 |
| Prior HF | 619 (3.8) | 552 (4.0) | 21 (3.6) | 46 (2.6) | 0.009 |
| Prior AF | 23 (0.1) | 21 (0.2) | 0 (0.0) | 2 (0.1) | 0.587 |
| Angina | 533 (3.3) | 425 (3.1) | 20 (3.4) | 88 (4.9) | <0.001 |
| Peripheral vascular<br>disease | 54 (0.3) | 48 (0.4) | 2 (0.3) | 4 (0.2) | 0.678 |
| Cerebrovascular<br>disease | 38 (0.2) | 35 (0.3) | 1 (0.2) | 2 (0.1) | 0.470 |
| Hypertension | 1355 (8.4) | 1177 (8.6) | 33 (5.6) | 145 (8.0) | 0.036 |
| **Other diagnoses** |  |  |  |  |  |
| Diabetes | 4648 (28.8) | 3979 (28.9) | 186 (31.6) | 483 (26.7) | 0.048 |
| Dementia | 135 (0.84) | 135 (1.0) | 0 (0.0) | 0 (0.0) | <0.001 |
| Cancer | 630 (3.9) | 589 (4.3) | 12 (2.0) | 29 (1.6) | <0.001 |
| Chronic pulmonary<br>disease | 881 (5.5) | 816 (5.9) | 16 (2.7) | 49 (2.7) | <0.001 |

MI Myocardial infarction; HF Heart failure; AF Atrial Fibrillation; CABG Coronary Artery Bypass Graft; PCI Percutaneous Coronary Intervention. \* p values refer to comparisons across the groups that did not receive cardiac rehabilitation, commenced but did not complete and completed cardiac rehabilitation and were tested by Kruskal Wallis test for continuous variables and Chi-square for categorical variables.

Median time from discharge to commencement of a cardiac rehabilitation program was 31 (IQR 18-51) days, respectively. Median duration of the cardiac rehabilitation program was 42 (IQR 30-61) days.

### Outcomes

#### Factors associated with non-completion

Living alone (OR 1.38; 95% CI 1.00-1.89; p=0.048), having diabetes (OR 1.48; 95% CI 1.02-2.13; p=0.037) or depression at baseline (OR 1.54; 95% CI 1.14-2.08; p=0.005), and being enrolled in the telephone program compared to a face-to-face program (OR 0.26; 95% CI 0.18-0.38; p<0.001) were associated with non-completion of cardiac rehabilitation.

#### Reasons for non-completion

Patient-related (not interested, n=109; feeling unwell, n=104; did not like or see value in the program, n= 9), service-related (preferring to follow-up with the general practitioner, n=46; cardiac rehabilitation program time unsuitable, n=8); and work/social circumstances-related (return to work, n=42; Transport/distance, n=34; cost, n=1) were reported as reasons for non-completion by 353 patients.

#### Clinical outcomes

There were 4,066 composite events of all-cause death, or admission for new/re-myocardial infarction, heart failure, atrial fibrillation or stroke) within the 12 months of follow-up after hospital discharge. After adjustment for sociodemographic and clinical factors, completing cardiac rehabilitation (HR 0.65; 95% CI 0.57-0.74; p<0.001) was associated with a lower risk of cardiovascular readmission/death. Commencing but not completing cardiac rehabilitation was not significantly associated with the composite outcome (HR 0.94; 95% CI 0.78-1.13; p=0.478)-Figure 3.

**Figure 3:**
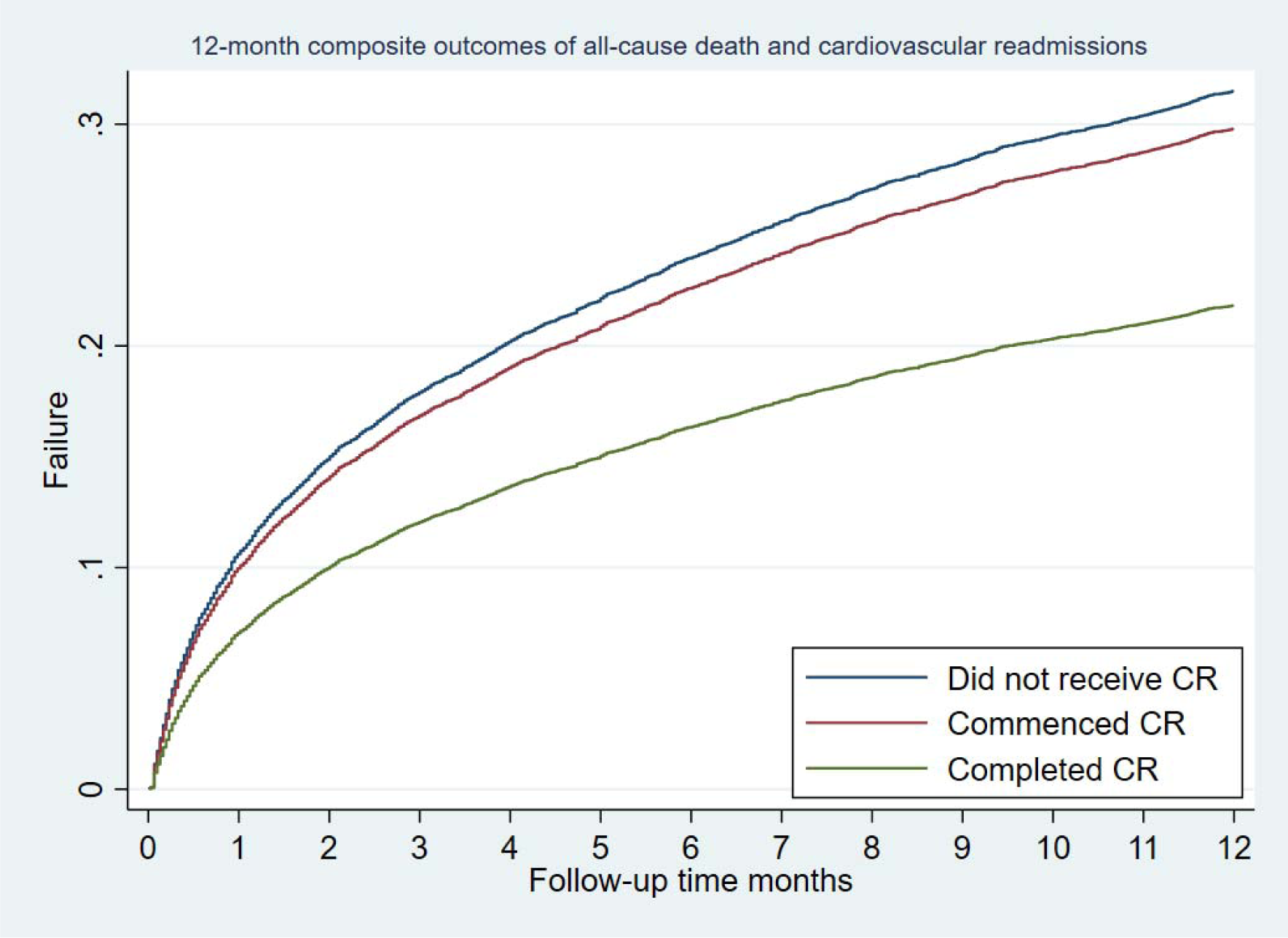
Adjusted all-cause mortality/cardiovascular readmission rates within 12 months after the index hospitalisation.

### Qualitative study

There were 28 participants in the qualitative study (24 participants from the five workshops focus groups interviews and 4 patients’ individual interviews). Participants’ median age was 66 (62-75) years and six (21.4%) were women. All had English as a first language, and none were of Aboriginal or Indigenous Australian heritage. One participant reported having a carer and two did not have access to the internet. Twelve (43%) were enrolled in a face-to-face program and 16 (57%) in the telephone program.

Five overarching themes (logistic barriers, social support, transition of care, program delivery and care integration) and 19 sub-themes emerged from this study (Table 2).

**Table 2.**
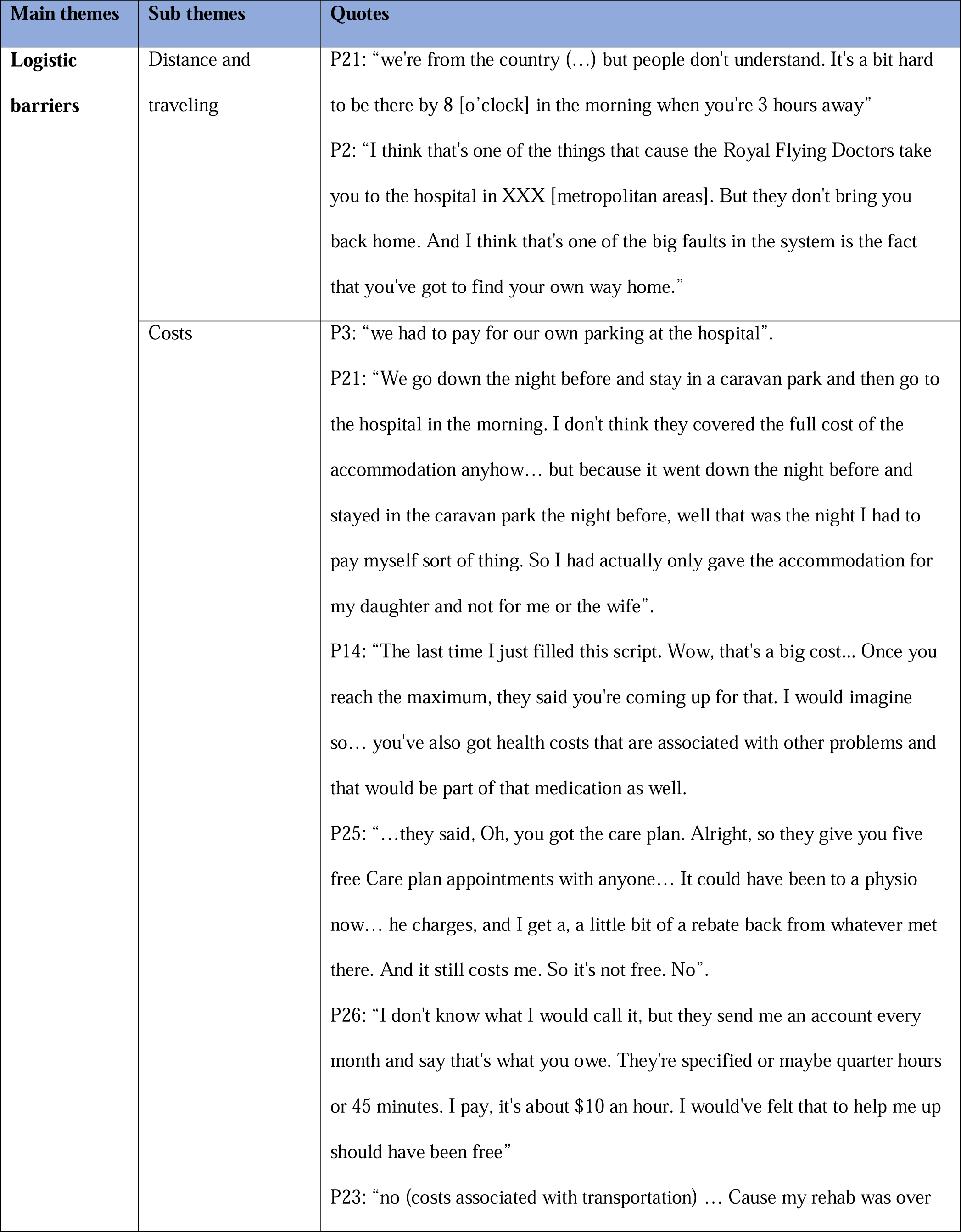
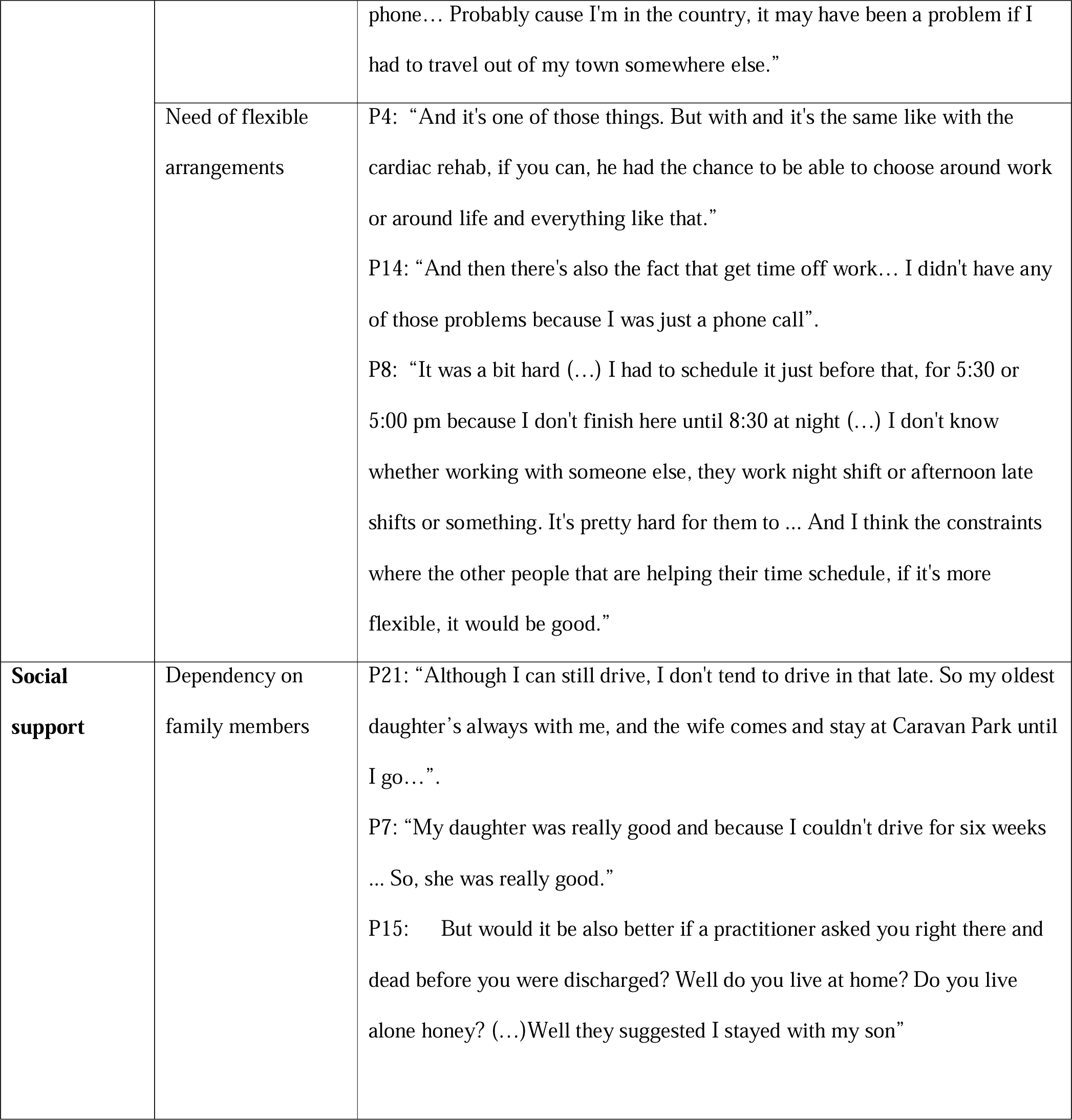
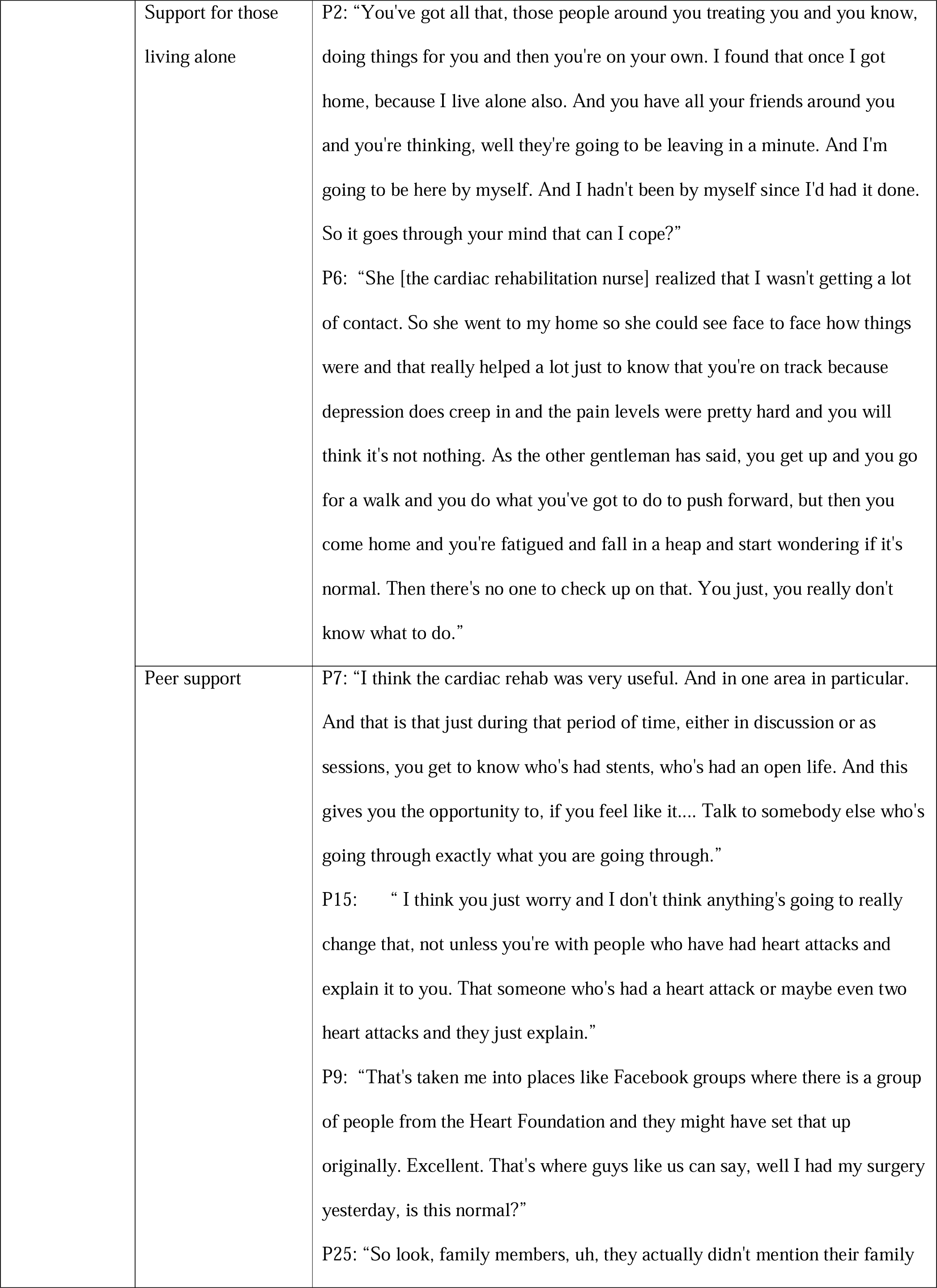
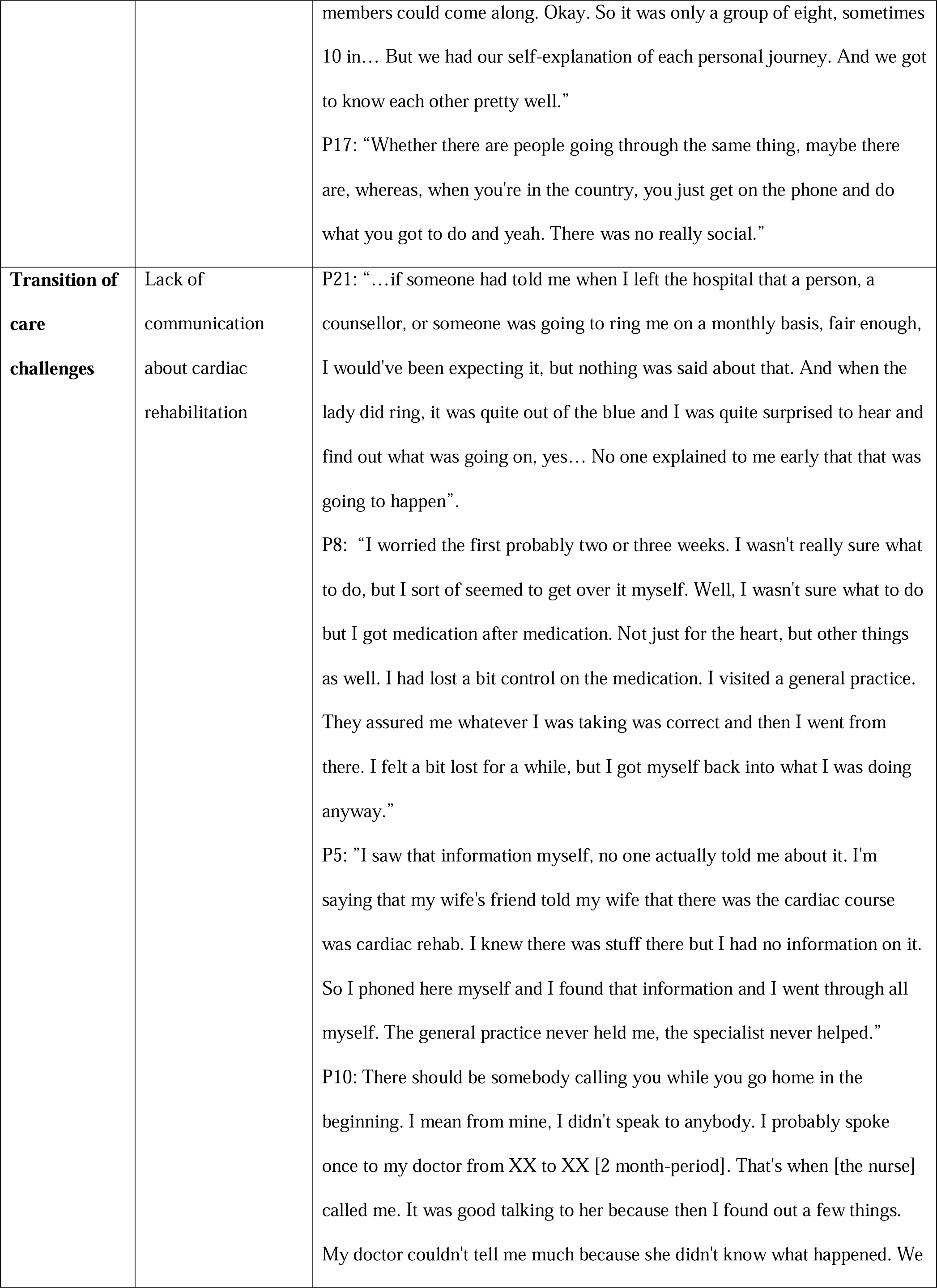
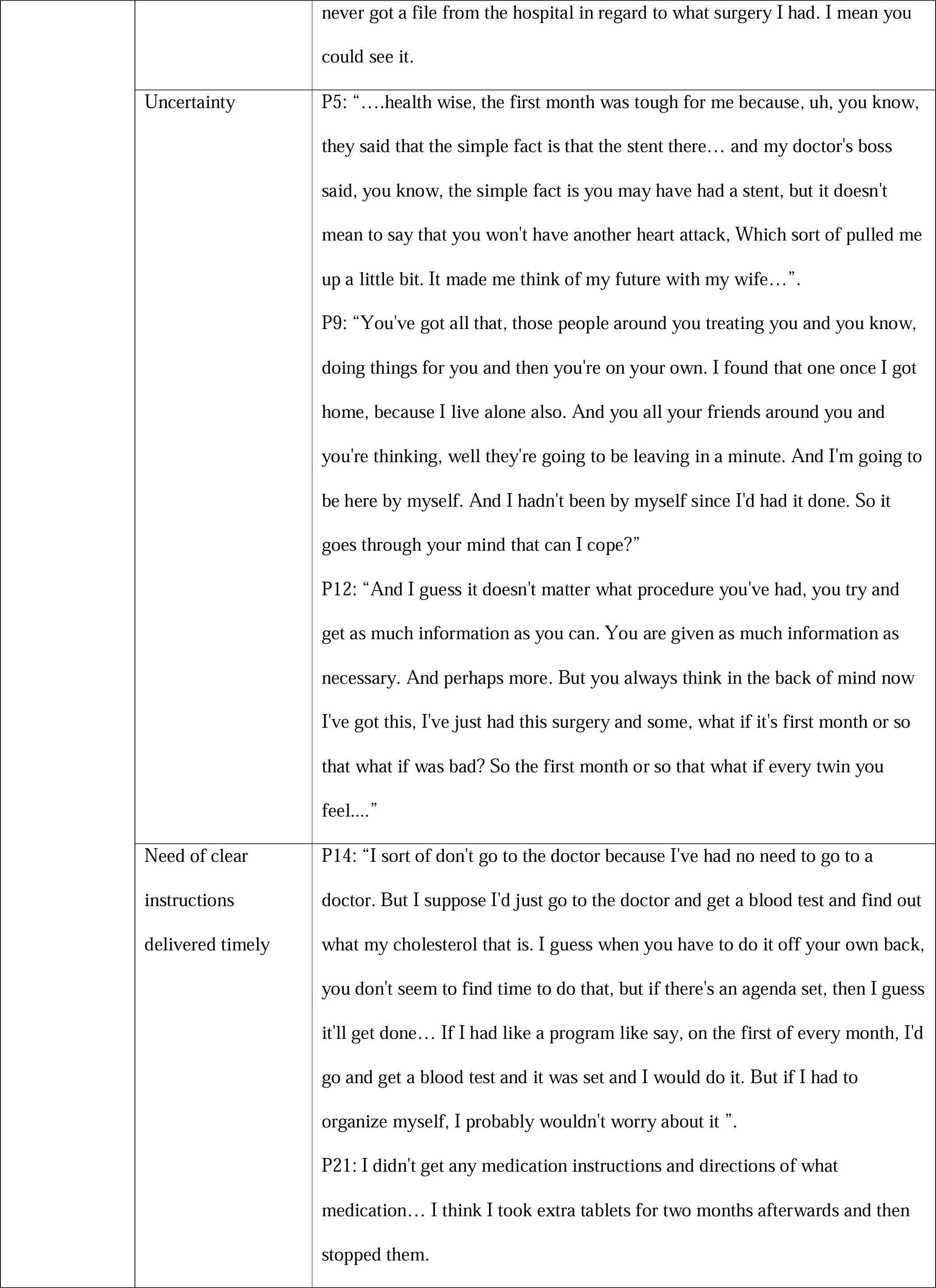
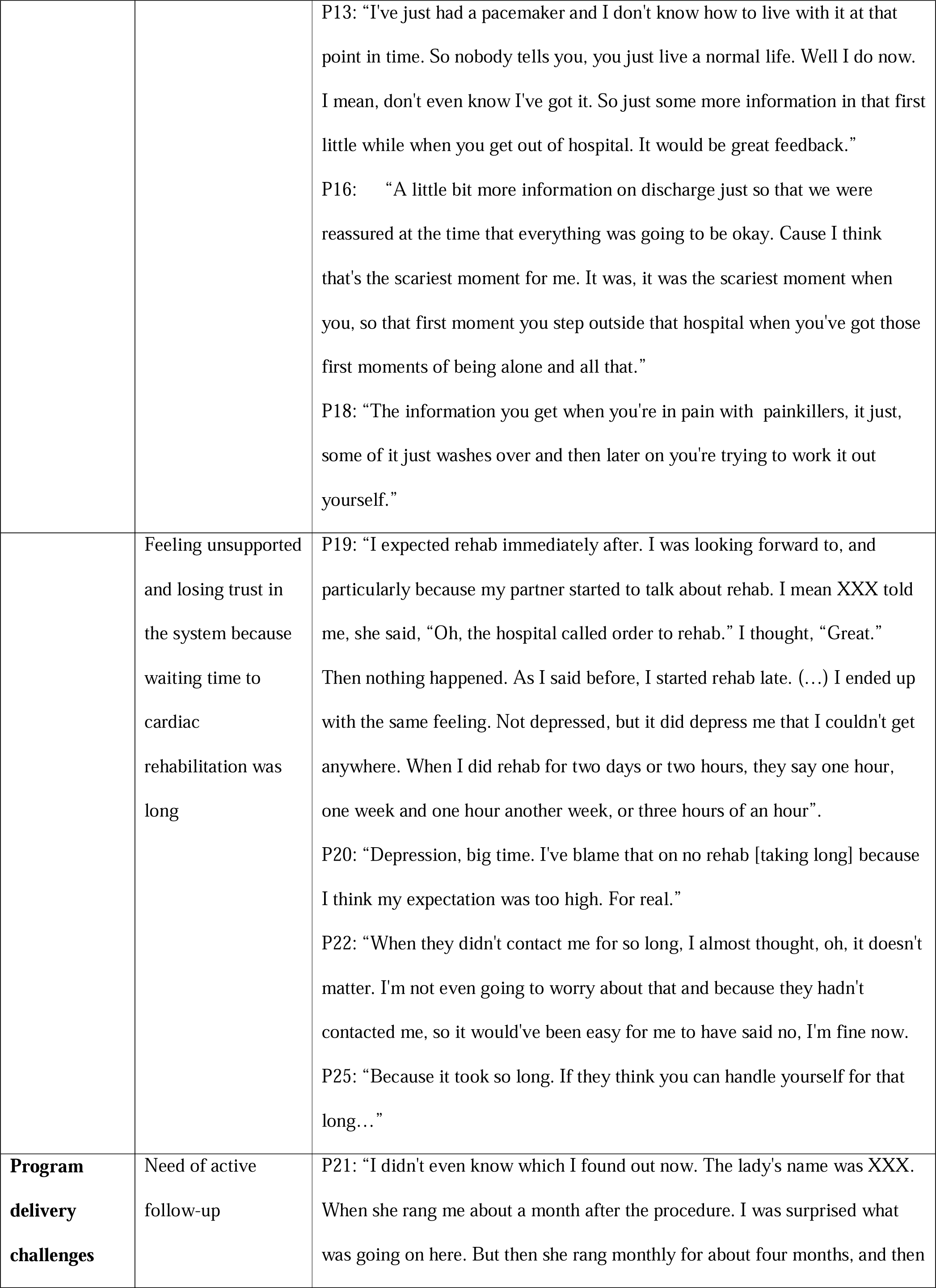
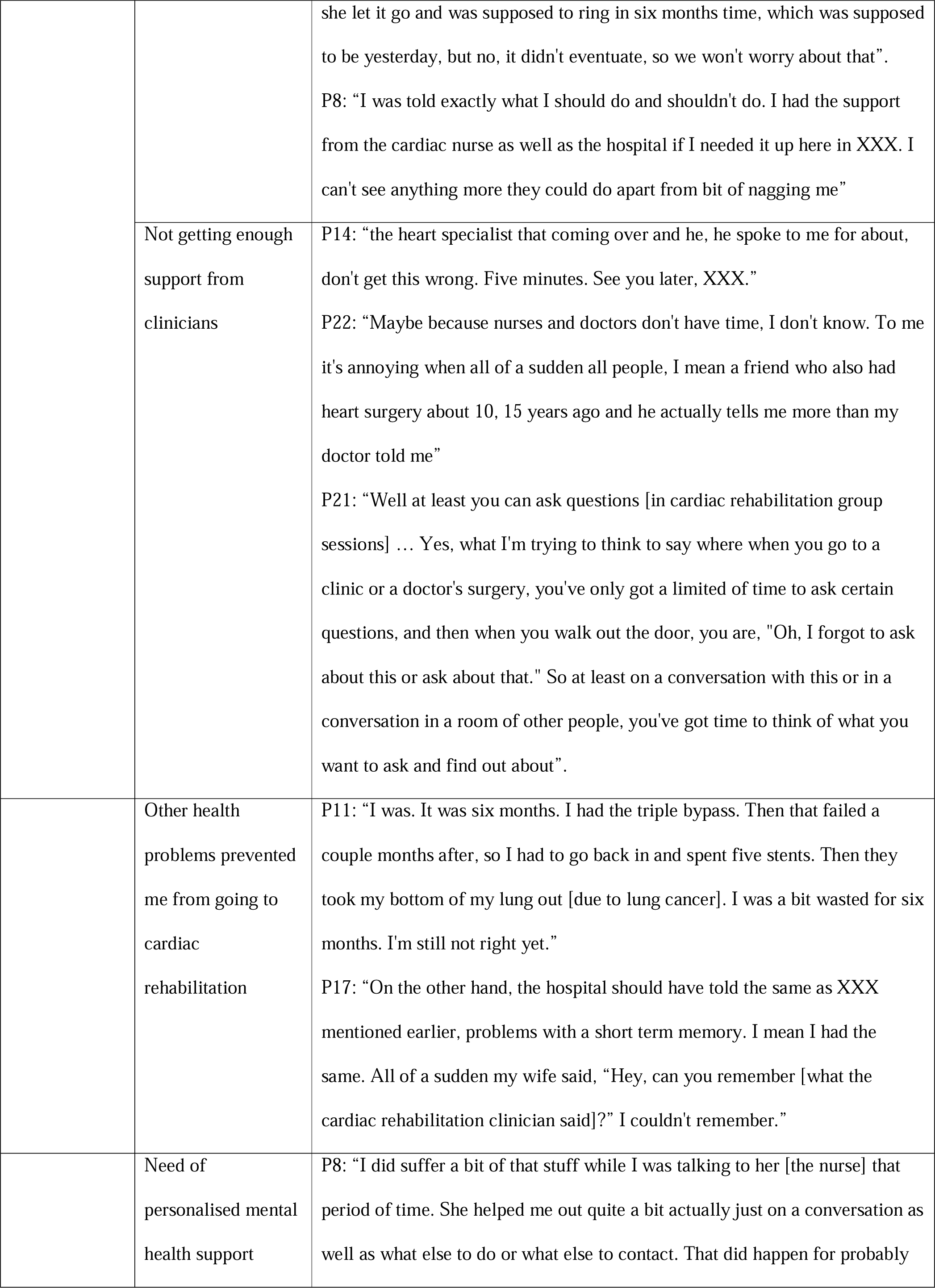
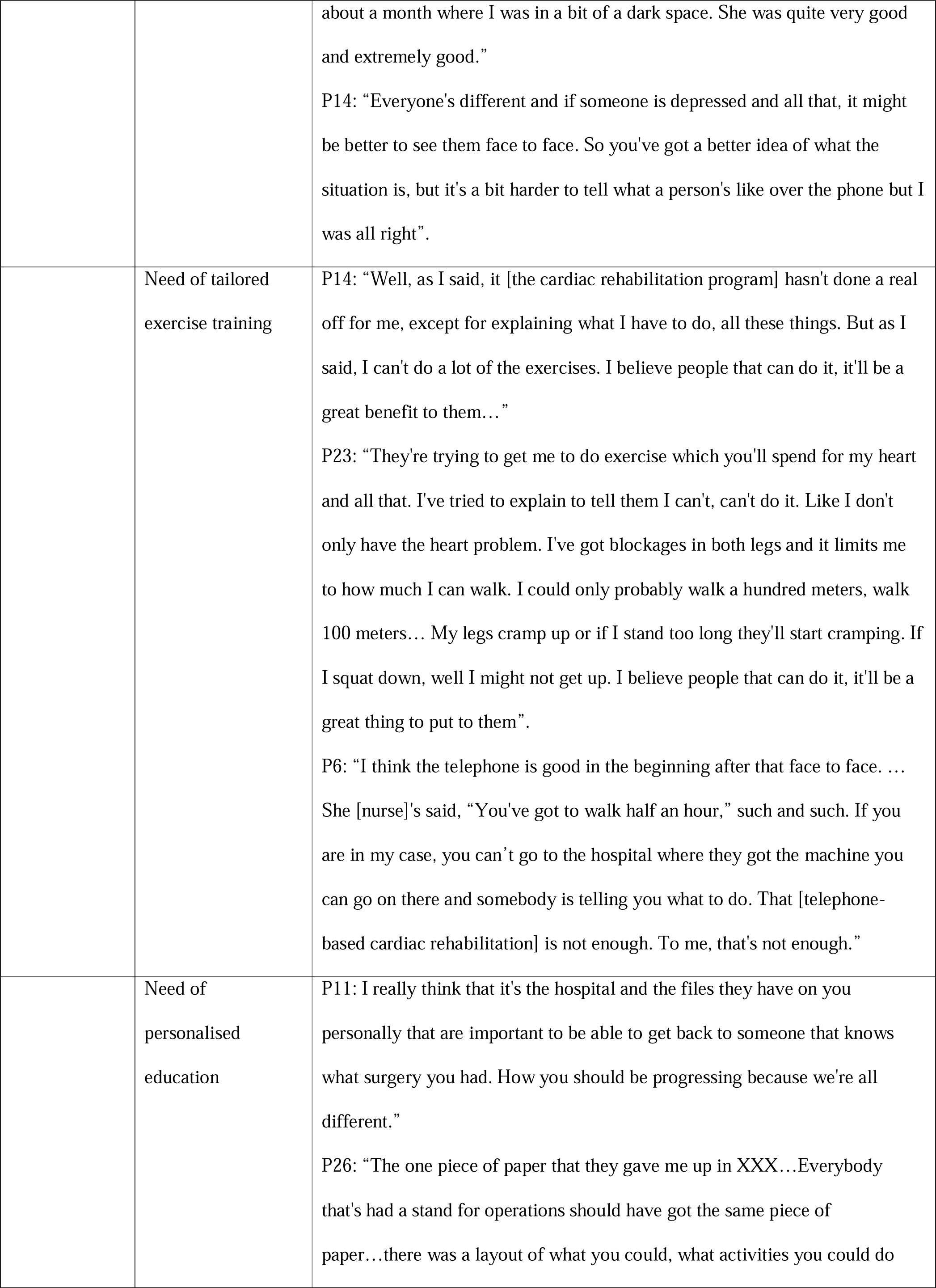
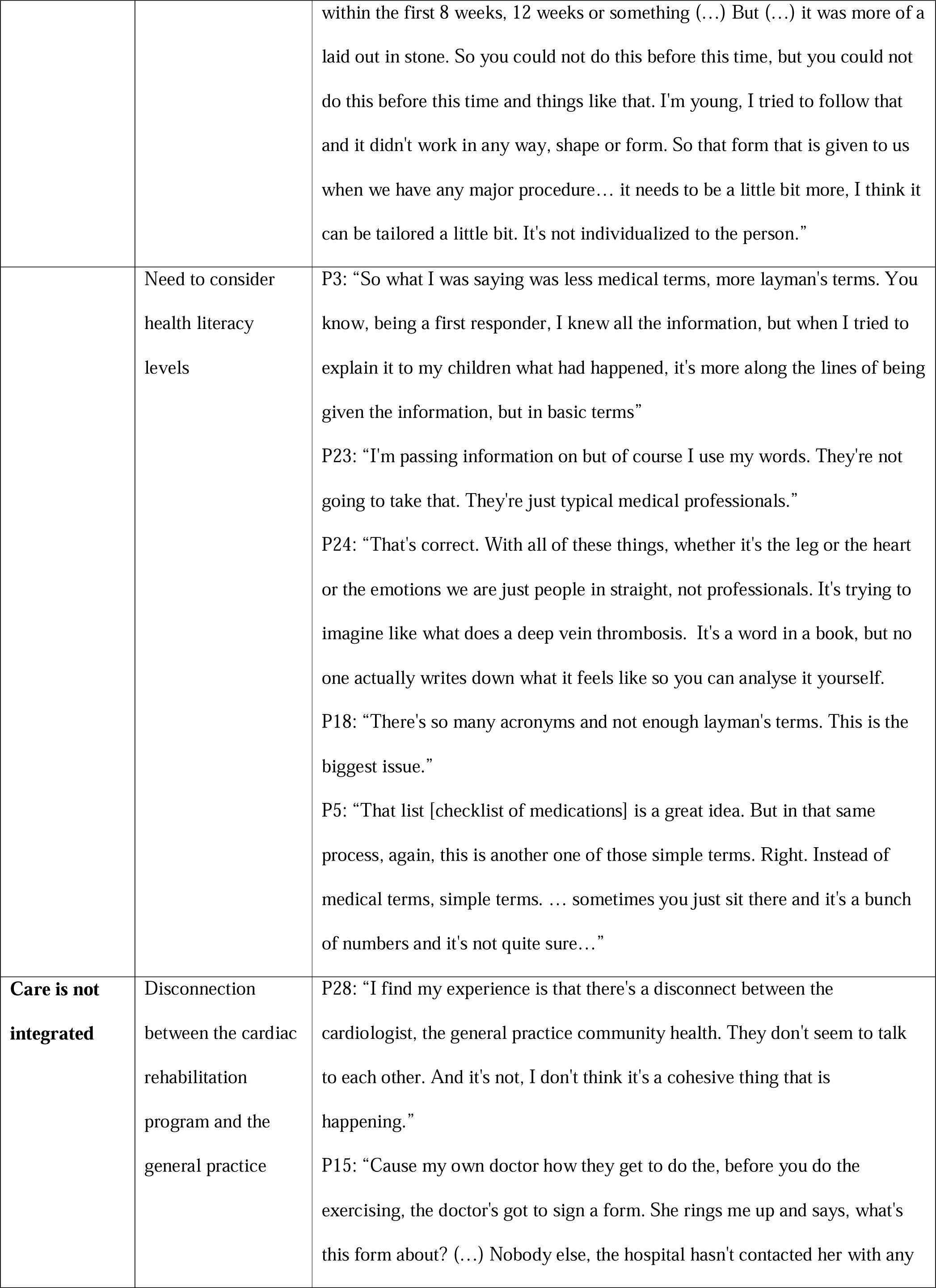
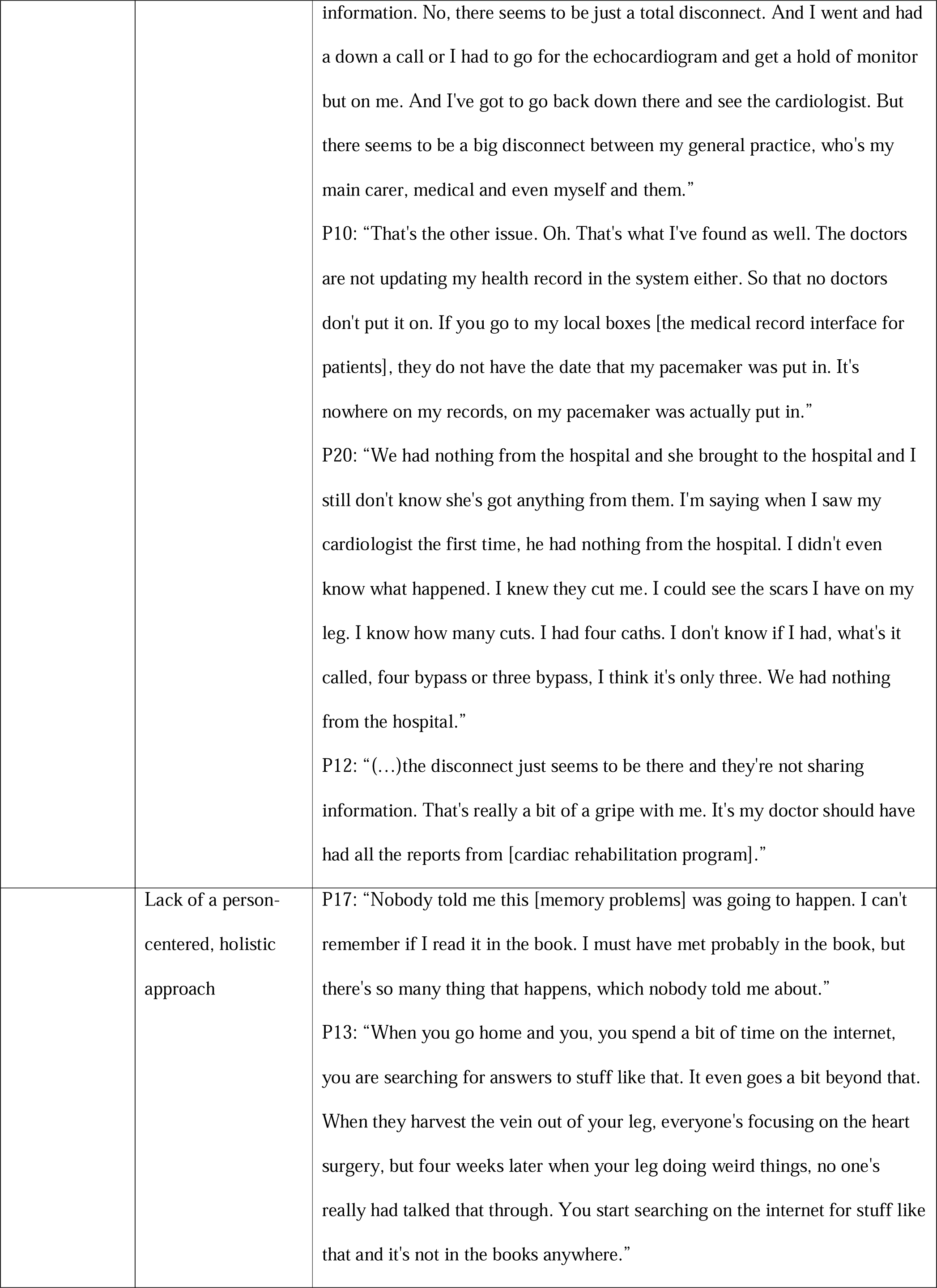
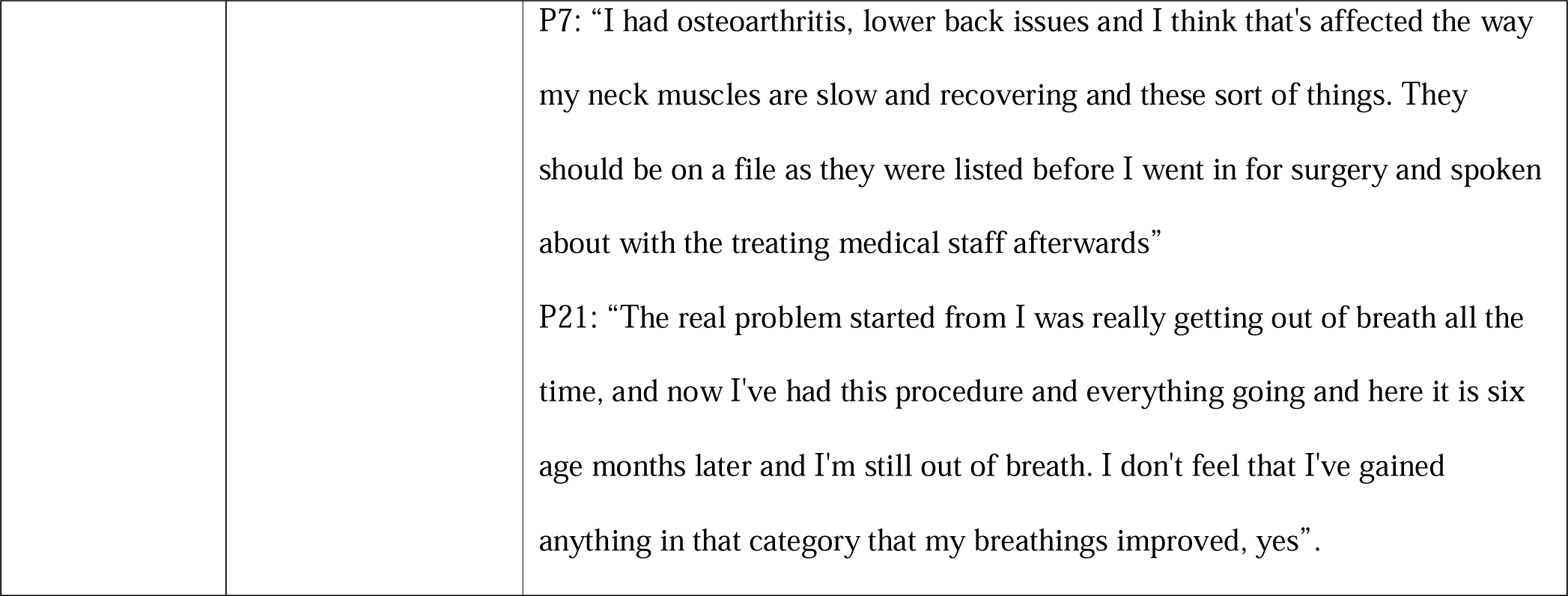
Themes and sub-themes that emerged from the qualitative study.

| Main themes | Sub themes | Quotes |
| --- | --- | --- |
| **Logistic barriers** | Distance and traveling | P21: “we're from the country (...) but people don't understand. It's a bit hard to be there by 8 [o'clock] in the morning when you're 3 hours away”<br>P2: “I think that's one of the things that cause the Royal Flying Doctors take you to the hospital in XXX [metropolitan areas]. But they don't bring you back home. And I think that's one of the big faults in the system is the fact that you've got to find your own way home.” |
| | Costs | P3: “we had to pay for our own parking at the hospital”.<br>P21: “We go down the night before and stay in a caravan park and then go to the hospital in the morning. I don't think they covered the full cost of the accommodation anyhow... but because it went down the night before and stayed in the caravan park the night before, well that was the night I had to pay myself sort of thing. So I had actually only gave the accommodation for my daughter and not for me or the wife”.<br>P14: “The last time I just filled this script. Wow, that's a big cost... Once you reach the maximum, they said you're coming up for that. I would imagine so... you've also got health costs that are associated with other problems and that would be part of that medication as well.<br>P25: “...they said, Oh, you got the care plan. Alright, so they give you five free Care plan appointments with anyone... It could have been to a physio now... he charges, and I get a, a little bit of a rebate back from whatever met there. And it still costs me. So it's not free. No”.<br>P26: “I don't know what I would call it, but they send me an account every month and say that's what you owe. They're specified or maybe quarter hours or 45 minutes. I pay, it's about \$10 an hour. I would've felt that to help me up should have been free”<br>P23: “no (costs associated with transportation) ... Cause my rehab was over |
|  |  | phone... Probably cause I'm in the country, it may have been a problem if I had to travel out of my town somewhere else.” |
|  | Need of flexible arrangements | P4: “And it's one of those things. But with and it's the same like with the cardiac rehab, if you can, he had the chance to be able to choose around work or around life and everything like that.”<br>P14: “And then there's also the fact that get time off work... I didn't have any of those problems because I was just a phone call”.<br>P8: “It was a bit hard (...) I had to schedule it just before that, for 5:30 or 5:00 pm because I don't finish here until 8:30 at night (...) I don't know whether working with someone else, they work night shift or afternoon late shifts or something. It's pretty hard for them to ... And I think the constraints where the other people that are helping their time schedule, if it's more flexible, it would be good.” |
| **Social support** | Dependency on family members | P21: “Although I can still drive, I don't tend to drive in that late. So my oldest daughter’s always with me, and the wife comes and stay at Caravan Park until I go...”.<br>P7: “My daughter was really good and because I couldn't drive for six weeks ... So, she was really good.”<br>P15: But would it be also better if a practitioner asked you right there and dead before you were discharged? Well do you live at home? Do you live alone honey? (...)Well they suggested I stayed with my son” |

|  |  |
| --- | --- |
| Support for those living alone | P2: “You've got all that, those people around you treating you and you know, doing things for you and then you're on your own. I found that once I got home, because I live alone also. And you have all your friends around you and you're thinking, well they're going to be leaving in a minute. And I'm going to be here by myself. And I hadn't been by myself since I'd had it done. So it goes through your mind that can I cope?”<br>P6: “She [the cardiac rehabilitation nurse] realized that I wasn't getting a lot of contact. So she went to my home so she could see face to face how things were and that really helped a lot just to know that you're on track because depression does creep in and the pain levels were pretty hard and you will think it's not nothing. As the other gentleman has said, you get up and you go for a walk and you do what you've got to do to push forward, but then you come home and you're fatigued and fall in a heap and start wondering if it's normal. Then there's no one to check up on that. You just, you really don't know what to do.” |
| Peer support | P7: “I think the cardiac rehab was very useful. And in one area in particular. And that is that just during that period of time, either in discussion or as sessions, you get to know who's had stents, who's had an open life. And this gives you the opportunity to, if you feel like it.... Talk to somebody else who's going through exactly what you are going through.”<br>P15: “ I think you just worry and I don't think anything's going to really change that, not unless you're with people who have had heart attacks and explain it to you. That someone who's had a heart attack or maybe even two heart attacks and they just explain.”<br>P9: “That's taken me into places like Facebook groups where there is a group of people from the Heart Foundation and they might have set that up originally. Excellent. That's where guys like us can say, well I had my surgery yesterday, is this normal?”<br>P25: “So look, family members, uh, they actually didn't mention their family |
|  |  | members could come along. Okay. So it was only a group of eight, sometimes 10 in... But we had our self-explanation of each personal journey. And we got to know each other pretty well.”<br>P17: “Whether there are people going through the same thing, maybe there are, whereas, when you're in the country, you just get on the phone and do what you got to do and yeah. There was no really social.” |
| **Transition of care challenges** | Lack of communication about cardiac rehabilitation | P21: “...if someone had told me when I left the hospital that a person, a counsellor, or someone was going to ring me on a monthly basis, fair enough, I would've been expecting it, but nothing was said about that. And when the lady did ring, it was quite out of the blue and I was quite surprised to hear and find out what was going on, yes... No one explained to me early that that was going to happen”.<br>P8: “I worried the first probably two or three weeks. I wasn't really sure what to do, but I sort of seemed to get over it myself. Well, I wasn't sure what to do but I got medication after medication. Not just for the heart, but other things as well. I had lost a bit control on the medication. I visited a general practice. They assured me whatever I was taking was correct and then I went from there. I felt a bit lost for a while, but I got myself back into what I was doing anyway.”<br>P5: “I saw that information myself, no one actually told me about it. I'm saying that my wife's friend told my wife that there was the cardiac course was cardiac rehab. I knew there was stuff there but I had no information on it. So I phoned here myself and I found that information and I went through all myself. The general practice never held me, the specialist never helped.”<br>P10: There should be somebody calling you while you go home in the beginning. I mean from mine, I didn't speak to anybody. I probably spoke once to my doctor from XX to XX [2 month-period]. That's when [the nurse] called me. It was good talking to her because then I found out a few things. My doctor couldn't tell me much because she didn't know what happened. We |
|  |  | never got a file from the hospital in regard to what surgery I had. I mean you could see it. |
|  | Uncertainty | P5: "...health wise, the first month was tough for me because, uh, you know, they said that the simple fact is that the stent there... and my doctor's boss said, you know, the simple fact is you may have had a stent, but it doesn't mean to say that you won't have another heart attack, Which sort of pulled me up a little bit. It made me think of my future with my wife...".<br>P9: "You've got all that, those people around you treating you and you know, doing things for you and then you're on your own. I found that one once I got home, because I live alone also. And you all your friends around you and you're thinking, well they're going to be leaving in a minute. And I'm going to be here by myself. And I hadn't been by myself since I'd had it done. So it goes through your mind that can I cope?"<br>P12: "And I guess it doesn't matter what procedure you've had, you try and get as much information as you can. You are given as much information as necessary. And perhaps more. But you always think in the back of mind now I've got this, I've just had this surgery and some, what if it's first month or so that what if was bad? So the first month or so that what if every twin you feel...." |
|  | Need of clear instructions delivered timely | P14: "I sort of don't go to the doctor because I've had no need to go to a doctor. But I suppose I'd just go to the doctor and get a blood test and find out what my cholesterol that is. I guess when you have to do it off your own back, you don't seem to find time to do that, but if there's an agenda set, then I guess it'll get done... If I had like a program like say, on the first of every month, I'd go and get a blood test and it was set and I would do it. But if I had to organize myself, I probably wouldn't worry about it".<br>P21: I didn't get any medication instructions and directions of what medication... I think I took extra tablets for two months afterwards and then stopped them. |
|  |  | P13: “I’ve just had a pacemaker and I don’t know how to live with it at that point in time. So nobody tells you, you just live a normal life. Well I do now. I mean, don’t even know I’ve got it. So just some more information in that first little while when you get out of hospital. It would be great feedback.”<br>P16: “A little bit more information on discharge just so that we were reassured at the time that everything was going to be okay. Cause I think that’s the scariest moment for me. It was, it was the scariest moment when you, so that first moment you step outside that hospital when you’ve got those first moments of being alone and all that.”<br>P18: “The information you get when you’re in pain with painkillers, it just, some of it just washes over and then later on you’re trying to work it out yourself.” |
|  | Feeling unsupported and losing trust in the system because waiting time to cardiac rehabilitation was long | P19: “I expected rehab immediately after. I was looking forward to, and particularly because my partner started to talk about rehab. I mean XXX told me, she said, “Oh, the hospital called order to rehab.” I thought, “Great.” Then nothing happened. As I said before, I started rehab late. (...) I ended up with the same feeling. Not depressed, but it did depress me that I couldn’t get anywhere. When I did rehab for two days or two hours, they say one hour, one week and one hour another week, or three hours of an hour”.<br>P20: “Depression, big time. I’ve blame that on no rehab [taking long] because I think my expectation was too high. For real.”<br>P22: “When they didn’t contact me for so long, I almost thought, oh, it doesn’t matter. I’m not even going to worry about that and because they hadn’t contacted me, so it would’ve been easy for me to have said no, I’m fine now.<br>P25: “Because it took so long. If they think you can handle yourself for that long...” |
| **Program delivery challenges** | Need of active follow-up | P21: “I didn’t even know which I found out now. The lady’s name was XXX. When she rang me about a month after the procedure. I was surprised what was going on here. But then she rang monthly for about four months, and then |
|  |  | she let it go and was supposed to ring in six months time, which was supposed to be yesterday, but no, it didn't eventuate, so we won't worry about that”.<br>P8: “I was told exactly what I should do and shouldn't do. I had the support from the cardiac nurse as well as the hospital if I needed it up here in XXX. I can't see anything more they could do apart from bit of nagging me” |
|  | Not getting enough support from clinicians | P14: “the heart specialist that coming over and he, he spoke to me for about, don't get this wrong. Five minutes. See you later, XXX.”<br>P22: “Maybe because nurses and doctors don't have time, I don't know. To me it's annoying when all of a sudden all people, I mean a friend who also had heart surgery about 10, 15 years ago and he actually tells me more than my doctor told me”<br>P21: “Well at least you can ask questions [in cardiac rehabilitation group sessions] ... Yes, what I'm trying to think to say where when you go to a clinic or a doctor's surgery, you've only got a limited of time to ask certain questions, and then when you walk out the door, you are, "Oh, I forgot to ask about this or ask about that." So at least on a conversation with this or in a conversation in a room of other people, you've got time to think of what you want to ask and find out about”. |
|  | Other health problems prevented me from going to cardiac rehabilitation | P11: “I was. It was six months. I had the triple bypass. Then that failed a couple months after, so I had to go back in and spent five stents. Then they took my bottom of my lung out [due to lung cancer]. I was a bit wasted for six months. I'm still not right yet.”<br>P17: “On the other hand, the hospital should have told the same as XXX mentioned earlier, problems with a short term memory. I mean I had the same. All of a sudden my wife said, “Hey, can you remember [what the cardiac rehabilitation clinician said]?” I couldn't remember.” |
|  | Need of personalised mental health support | P8: “I did suffer a bit of that stuff while I was talking to her [the nurse] that period of time. She helped me out quite a bit actually just on a conversation as well as what else to do or what else to contact. That did happen for probably |
|  |  | about a month where I was in a bit of a dark space. She was quite very good and extremely good.”<br>P14: “Everyone's different and if someone is depressed and all that, it might be better to see them face to face. So you've got a better idea of what the situation is, but it's a bit harder to tell what a person's like over the phone but I was all right”. |
|  | Need of tailored exercise training | P14: “Well, as I said, it [the cardiac rehabilitation program] hasn't done a real off for me, except for explaining what I have to do, all these things. But as I said, I can't do a lot of the exercises. I believe people that can do it, it'll be a great benefit to them...”<br>P23: “They're trying to get me to do exercise which you'll spend for my heart and all that. I've tried to explain to tell them I can't, can't do it. Like I don't only have the heart problem. I've got blockages in both legs and it limits me to how much I can walk. I could only probably walk a hundred meters, walk 100 meters... My legs cramp up or if I stand too long they'll start cramping. If I squat down, well I might not get up. I believe people that can do it, it'll be a great thing to put to them”.<br>P6: “I think the telephone is good in the beginning after that face to face. ... She [nurse]'s said, “You've got to walk half an hour,” such and such. If you are in my case, you can't go to the hospital where they got the machine you can go on there and somebody is telling you what to do. That [telephone-based cardiac rehabilitation] is not enough. To me, that's not enough.” |
|  | Need of personalised education | P11: I really think that it's the hospital and the files they have on you personally that are important to be able to get back to someone that knows what surgery you had. How you should be progressing because we're all different.”<br>P26: “The one piece of paper that they gave me up in XXX...Everybody that's had a stand for operations should have got the same piece of paper...there was a layout of what you could, what activities you could do |
|  |  | within the first 8 weeks, 12 weeks or something (...) But (...) it was more of a laid out in stone. So you could not do this before this time, but you could not do this before this time and things like that. I'm young, I tried to follow that and it didn't work in any way, shape or form. So that form that is given to us when we have any major procedure... it needs to be a little bit more, I think it can be tailored a little bit. It's not individualized to the person.” |
|  | Need to consider health literacy levels | P3: “So what I was saying was less medical terms, more layman's terms. You know, being a first responder, I knew all the information, but when I tried to explain it to my children what had happened, it's more along the lines of being given the information, but in basic terms”<br>P23: “I'm passing information on but of course I use my words. They're not going to take that. They're just typical medical professionals.”<br>P24: “That's correct. With all of these things, whether it's the leg or the heart or the emotions we are just people in straight, not professionals. It's trying to imagine like what does a deep vein thrombosis. It's a word in a book, but no one actually writes down what it feels like so you can analyse it yourself.<br>P18: “There's so many acronyms and not enough layman's terms. This is the biggest issue.”<br>P5: “That list [checklist of medications] is a great idea. But in that same process, again, this is another one of those simple terms. Right. Instead of medical terms, simple terms. ... sometimes you just sit there and it's a bunch of numbers and it's not quite sure...” |
| **Care is not integrated** | Disconnection between the cardiac rehabilitation program and the general practice | P28: “I find my experience is that there's a disconnect between the cardiologist, the general practice community health. They don't seem to talk to each other. And it's not, I don't think it's a cohesive thing that is happening.”<br>P15: “Cause my own doctor how they get to do the, before you do the exercising, the doctor's got to sign a form. She rings me up and says, what's this form about? (...) Nobody else, the hospital hasn't contacted her with any |
|  |  | information. No, there seems to be just a total disconnect. And I went and had a down a call or I had to go for the echocardiogram and get a hold of monitor but on me. And I've got to go back down there and see the cardiologist. But there seems to be a big disconnect between my general practice, who's my main carer, medical and even myself and them.”<br>P10: “That's the other issue. Oh. That's what I've found as well. The doctors are not updating my health record in the system either. So that no doctors don't put it on. If you go to my local boxes [the medical record interface for patients], they do not have the date that my pacemaker was put in. It's nowhere on my records, on my pacemaker was actually put in.”<br>P20: “We had nothing from the hospital and she brought to the hospital and I still don't know she's got anything from them. I'm saying when I saw my cardiologist the first time, he had nothing from the hospital. I didn't even know what happened. I knew they cut me. I could see the scars I have on my leg. I know how many cuts. I had four caths. I don't know if I had, what's it called, four bypass or three bypass, I think it's only three. We had nothing from the hospital.”<br>P12: “(...)the disconnect just seems to be there and they're not sharing information. That's really a bit of a gripe with me. It's my doctor should have had all the reports from [cardiac rehabilitation program].” |
|  | Lack of a person-centered, holistic approach | P17: “Nobody told me this [memory problems] was going to happen. I can't remember if I read it in the book. I must have met probably in the book, but there's so many thing that happens, which nobody told me about.”<br>P13: “When you go home and you, you spend a bit of time on the internet, you are searching for answers to stuff like that. It even goes a bit beyond that. When they harvest the vein out of your leg, everyone's focusing on the heart surgery, but four weeks later when your leg doing weird things, no one's really had talked that through. You start searching on the internet for stuff like that and it's not in the books anywhere.” |
|  |  | P7: “I had osteoarthritis, lower back issues and I think that's affected the way my neck muscles are slow and recovering and these sort of things. They should be on a file as they were listed before I went in for surgery and spoken about with the treating medical staff afterwards”<br>P21: “The real problem started from I was really getting out of breath all the time, and now I've had this procedure and everything going and here it is six age months later and I'm still out of breath. I don't feel that I've gained anything in that category that my breathings improved, yes”. |

### Convergence of qualitative and quantitative findings

We found convergence between the factors and reasons found in the quantitative analysis and the barriers and needs that emerged in the qualitative study (Table 3). However, for telehealth, which was associated with lower chance of non-completion (i.e., telehealth was associated with higher completion), we found some divergence between the themes. Whereas the theme “logistic barriers” converged with the findings of telehealth increasing cardiac rehabilitation participation, in the qualitative study the emerging sub-themes “need of personalized mental health support”, “need of tailored exercise training” and “(lack) peer support” were reported as barriers associated with telehealth.

**Table 3:** Convergence of factors and reasons associated with non-completion and themes and sub-themes emerging from the qualitative study.

| Factor/Reason | Theme | Subtheme | Exemplar quotes |
| --- | --- | --- | --- |
| Depression | Transition of care challenges | Feeling unsupported and losing trust in the system because waiting time to cardiac rehabilitation was long | P20: "Depression, big time. I've blamed that on no rehab because I think my expectation was too high. For real."<br>P19: "I expected rehab immediately after. I was looking forward to, and particularly because my partner started to talk about rehab. I mean XXX told me, she said, "Oh, the hospital called order to rehab." I thought, "Great." Then nothing happened. As I said before, I started rehab late. (...) I ended up with the same feeling. Not depressed, but it did depress me that I couldn't get anywhere. When I did rehab for two days or two hours, they say one hour, one week and one hour another week, or three hours of an hour". |
|  |  | Need of tailored mental health support | P14: "Everyone's different and if someone is depressed and all that, it might be better to see them face to face. So you've got a better idea of what the situation is."<br>P8: "I did suffer a bit of that stuff while I was talking to her [the nurse] that period of time. She helped me out quite a bit actually just on a conversation as well as what else to do or what else to contact. That did happen for probably about a month where I was in a bit of a dark space. She was quite very good and extremely good." |
|  |  | Support for those living alone | P6: "She [the cardiac rehabilitation nurse] realized that I wasn't getting a lot of contact. So she went to my home so she could see face to face how things were and that really helped a lot just to know that you're on track because depression does creep in and the pain levels were pretty hard and you will think it's not nothing. As the other gentleman has said, you get up and you go for a walk and you do what you've got to do to push forward, but then you come home and you're fatigued and fall in a heap and start wondering if it's normal. Then there's no one to check up on that. You just, you really don't know what to do." |
| Living alone | Transition of care challenges | Uncertainty | P9: "You've got all that, those people around you treating you and you know, doing things for you and then you're on your own. I found that one once I got home, because I live alone also. And you all your friends around you and you're thinking, well they're going to be leaving in a minute. And I'm going to be here by myself. And I hadn't been by myself since I'd had it done. So it goes through your mind that can I cope?" |
|  | Social support | Dependency on family members | P15: "But would it be also better if a practitioner asked you right there and dead before you were discharged. Well do you live at home? Do you live alone honey? (...)Well they suggested I stayed with my son" |
| Telehealth | Logistic barriers | Distance and travel | P21: "we're from the country (...) but people don't understand. It's a bit hard to be there by 8 [o'clock] in the morning when you're 3 hours away" |
|  | Logistic barriers | Flexible arrangements | P14: "And then there's also the fact that get time off work... I didn't have any of those problems because I was just a phone call". |
|  | Logistic barriers | Costs | P23: "no [costs associated with transportation] ... Cause my rehab was over phone... Probably cause I'm in the country, it may have been a problem if I had to travel out of my town somewhere else.” |
|  | Program delivery challenges | * Need of personalised mental health support | P14: “Everyone's different and if someone is depressed and all that, it might be better to see them face to face. So you've got a better idea of what the situation is, but it's a bit harder to tell what a person's like over the phone but I was all right”. |
|  | Program delivery challenges | * Need of tailored exercise training | P6: “I think the telephone is good in the beginning after that face to face. ... She [nurse]'s said, “You've got to walk half an hour,” such and such. If you are in my case, you can't go to the hospital where they got the machine you can go on there and somebody is telling you what to do. That [telephone-based cardiac rehabilitation] is not enough. To me, that's not enough.” |
|  | Social support | *Peer support | P17: “Whether there are people going through the same thing, maybe there are, whereas, when you're in the country, you just get on the phone and do what you got to do and yeah. There was no really social.” |
| Not interested | Transition of care challenges | Lack of communication about cardiac rehabilitation | P21: “...if someone had told me when I left the hospital that a person, a counsellor, or someone was going to ring me on a monthly basis, fair enough, I would've been expecting it, but nothing was said about that. And when the lady did ring, it was quite out of the blue and I was quite surprised to hear and find out what was going on, yes... No one explained to me early that that was going to happen”. |
|  | Transition of care challenges | Need of clear instructions delivered timely | P14: “I sort of don't go to the doctor because I've had no need to go to a doctor. But I suppose I'd just go to the doctor and get a blood test and find out what my cholesterol that is. I guess when you have to do it off your own back, you don't seem to find time to do that, but if there's an agenda set, then I guess it'll get done... If I had like a program like say, on the first of every month, I'd go and get a blood test and it was set and I would do it. But if I had to organize myself, I probably wouldn't worry about it ”. |
|  | Program delivery challenges | Need of active follow-up | P21: “I didn't even know which I found out now. The lady's name was XXX. When she rang me about a month after the procedure. I was surprised what was going on here. But then she rang monthly for about four months, and then she let it go and was supposed to ring in six months time, which was supposed to be yesterday, but no, it didn't eventuate, so we won't worry about that”. |
|  | Program delivery challenges | Need to consider health literacy levels | P18: “There's so many acronyms and not enough layman's terms. This is the biggest issue.”<br>P5: “That list [checklist of medications] is a great idea. But in that same process, again, this is another one of those simple terms. Right. Instead of medical terms, simple terms. ... sometimes you just sit there and it's a bunch of numbers and it's not quite sure...” |
|  | Transition of care challenges | Feeling unsupported and losing trust in the system because waiting time to cardiac rehabilitation was long | P22: “When they didn't contact me for so long, I almost thought, oh, it doesn't matter. I'm not even going to worry about that and because they hadn't contacted me, so it would've been easy for me to have said no, I'm fine now.<br>P25: “Because it took so long. If they think you can handle yourself for that long...” |
| Preferring to follow-up with | Care is not integrated | Disconnection between the | P28: “I find my experience is that there's a disconnect between the cardiologist, the general practice community |
| the general practitioner |  | cardiac rehabilitation program and the general practice | health. They don't seem to talk to each other. And it's not, I don't think it's a cohesive thing that is happening.”<br>P15: “Cause my own doctor how they get to do the, before you do the exercising, the doctor's got to sign a form. She rings me up and says, what's this form about? (...) Nobody else, the hospital hasn't contacted her with any information. No, there seems to be just a total disconnect. And I went and had a down a call or I had to go for the echocardiogram and get a hold of monitor but on me. And I've got to go back down there and see the cardiologist. But there seems to be a big disconnect between my general practice, who's my main carer, medical and even myself and them.”<br>P12: “(...) the disconnect just seems to be there and they're not sharing information. That's really a bit of a gripe with me. It's my doctor should have had all the reports from [cardiac rehabilitation program].” |
|  |  | Not getting enough support from clinicians | P14: “the heart specialist that coming over and he, he spoke to me for about, don't get this wrong. Five minutes. See you later, XXX.” |

## Discussion

This study showed low utilisation rates of cardiac rehabilitation by people of low socioeconomic status living in rural Australia. Whereas completing the program was associated with a 35% reduction in the risk of death or hospital admission, attending the program without completing it was not associated with a risk reduction. The main reasons for non-completion were lack of interest, being unwell and preferring to follow-up within primary care only. While living alone and having diabetes or depression increased the risk of non-completion, being enrolled in a telehealth program decreased the risk of not completing. There was convergence of the factors/reasons with the themes emerging from the qualitative study except for needs around telehealth-delivered cardiac rehabilitation.

Lemstra et al. (2013) showed lower cardiac rehabilitation attendance and completion rates (6.4% versus 14.8% in our study and 37.9% versus 74.6%, respectively) and higher waiting times (123 days vs 31 days) among people living in low-income neighbourhoods in Canada after a hospitalisation for ischemic heart disease or revascularisation procedure.^11^ Conversely, completion rates among those who started cardiac rehabilitation were higher in a Danish study than in ours (97.7% vs 74.6%).^22^ Their model of care included an extra number of cardiac rehabilitation sessions, telephone follow-up and involvement of general practices and patient associations in the maintenance phase for their socially vulnerable group.

Living alone was associated with a higher risk of not completing cardiac rehabilitation by our quantitative study and reported as a barrier to cardiac rehabilitation participation in our qualitative study. Living alone was related to feelings of being unsupported which coincide with findings that it is a proxy of low social support and worse clinical outcomes.^23^

Long waiting times were reported as a contributing factor to feelings depressed and unsupported leading to a decreased interest in cardiac rehabilitation. Likewise, lack of or unclear communication about cardiac rehabilitation during the hospital stay and transition of care period were referred as sources of low interest in cardiac rehabilitation which relates to evidence that physician endorsement^24^ and enrolment by a nurse or allied health professional increases cardiac rehabilitation participation.^10^

Another factor that could be leading to disinterest in cardiac rehabilitation is clinician communication occurring at a health literacy level that was not appropriate as reported in our qualitative study. As health literacy may influence both patient satisfaction and self-management of risk factors, ^25^ programs should consider evaluating health literacy and addressing the patient’s needs. This may involve helping patients with (*i*) finding and (*ii*) appraising health information, (*iii*) navigating the health care system, (*iv*) engaging with health care providers.^26^

The perception that the general practitioner was not involved with cardiac rehabilitation may reduce the program value to patients leading to low participation. Ensuring that an intake and discharge transition records are shared with the primary care provider can improve integration of care. However, <50% of the general practices receive an intake transition record from the cardiac rehabilitation program.^27^

Cardiac rehabilitation delivered via telehealth was independently associated with cardiac rehabilitation completion, which can be explained by telehealth increasing access to care by minimising logistic barriers such as distance and travelling, costs and scheduling flexibility.^28^ However, participants highlighted that mental health support, exercise training and peer support were barriers to completion faced in the telehealth program. Tailoring of the psychosocial components of cardiac rehabilitation to women’s needs has been associated with gains in health-related quality of life when cardiac rehabilitation.^29^ Moreover, offering alternative forms of exercise, such as yoga and Tai Chi Chuan, could facilitate personalisation and lead to improvements in cardiac rehabilitation adherence and participation particularly among women.^30^

As limitations of our study, we acknowledge that its observational design limits generalisability and that, despite the use of the inverse probability weighting and extensive model adjustment, selection bias and residual confounding may still have occurred despite adjustment for clinical and sociodemographic factors. Additionally, it is likely that women, people with English as a second language and Aboriginal and Torrens Strait Islander peoples were underrepresented in the qualitative study which may have impacted the emergence of the themes.

As messages to clinical practice, we would highlight that, as program completion was key to reduce the risk of hospitalisations and death among people of low socioeconomic status living in rural areas, it would be advisable to concentrate cardiac rehabilitation quality improvement initiatives around issues that may lead to improved completion. These may include: (i) identifying patients who live alone and providing them with social support; (ii) reducing waiting time; (iii) communicating the benefits of cardiac rehabilitation timely and at an appropriate health literacy level; (iv) integrating cardiac rehabilitation with primary care; (iv) offering telehealth as an option of cardiac rehabilitation delivery and designing telehealth models that enable tailored mental health support, exercise training and peer support; (v) personalising mental health support and exercise program.

In conclusion, our findings reinforce that completion of cardiac rehabilitation is paramount to achievement of clinical benefits in patients with low socioeconomic status living in rural areas. However, only 11.2% of the eligible patients are completing cardiac rehabilitation. Addressing barriers and needs related to logistics, social support particularly to those living alone, transition of care, care integration and lack of person-centredness of the programs especially through telehealth-enabled models of care may increase the interest and value of patients with socioeconomic disadvantaged leading to improved participation and clinical outcomes.

## Clinical Messages

- Program completion reduces the risk of hospitalisation and death among people of low socioeconomic status.
- Focus cardiac rehabilitation initiatives on:
21.
i. identifying and supporting patients living alone
ii. reducing waiting time
iii. offering telehealth as an option of program delivery, personalising, and improving mental health support, exercise, and peer support

## Data Availability

All data produced in the present study are available upon reasonable request to the authors

## Acknowledgments

We acknowledge the contribution of the cardiac rehabilitation patients, South Australian public health cardiac rehabilitation service providers and the Heads of Unit across the Department of Health Local Health Networks, the SA/NT Datalink consortium for providing the data linkage and the Integrated Cardiovascular Clinical Network, Rural Support Service, South Australia.

## Authors’ contributions

The study was conceived and designed by AB and RC. AB, MAPP, CH, RC, WK, ML, PT, and JH secured funds for the study. AB analysed the quantitative data. AB and HD analysed the qualitative data with input from MAPP and CH. JSR, OS and TM collected the qualitative data. The first draft of the paper was written by AB with input from all co-authors. All authors read and approved the final manuscript.

## Data availability statement

The data underlying this article cannot be shared publicly due to SA Health policy for data linkage privacy. The data will be shared on reasonable request to the corresponding author.

